# Neighborhood deprivation is associated with NICU mortality for extremely premature infants: A 4-NICU study

**DOI:** 10.1101/2022.09.28.22280470

**Authors:** Brynne A. Sullivan, Ayush Doshi, Pavel Chernyavskiy, Ameena Husain, Alexandra Binai, Rakesh Sahni, Karen D. Fairchild, J. Randall Moorman, Colm P. Travers, Zachary A. Vesoulis

## Abstract

**Importance:** Socioeconomic status impacts pregnancy outcomes and child development after NICU discharge for infants born prematurely, but has not been well studied for outcomes during the NICU stay. The Area Deprivation Index (ADI) is a validated measure of neighborhood disadvantage that uses Census data on income, education, employment, and housing quality.

**Objective:** In NICUs in different US regions, determine if ADI predicts NICU mortality and morbidity in extremely premature infants.

**Design:** We conducted a retrospective cohort study.

**Setting:** Four level IV neonatal intensive care units (NICU) in different US geographic regions: Northeast, Mid-Atlantic, Midwest, and South.

**Participants:** Non-Hispanic White and Black extremely premature infants (gestational age <29 weeks) and admitted to a study NICU from 2012-2020.

**Exposures:** ADI, race, BW, sex, and outborn status (admitted after transfer from an outside birth hospital).

**Main Outcomes and Measures:** We converted addresses to census blocks, identified by 12-digit Federal Information Processing Series (FIPS) codes, to link residences to the national ADI percentile of study participants. We analyzed the relationship between ADI and NICU mortality using Bayesian logistic regression adjusted for race, BW, outborn status, and sex. Predictors were considered significant if the 95% Credible Intervals excluded zero. We also analyzed the effect of ADI on NICU morbidities of late-onset sepsis, necrotizing enterocolitis, and severe intraventricular hemorrhage.

**Results:** We studied 2,765 infants. In univariate analysis, infants with higher ADI were more likely to be Black, suffer from short-term morbidities, and die before NICU discharge. ADI did not correlate with BW (r = −0.05) or sex. Black infants also had higher mortality and lower BW. In a multivariable model, lower BW, higher ADI, and male sex were statistically significant risk factors, while Black race and outborn status were not. Using these methods, ADI was also identified as a risk factor for NICU morbidities.

**Conclusions and Relevance:** Among extremely preterm infants admitted to four NICUs in different US geographic regions, ADI was a risk factor for mortality and morbidity after adjusting for multiple covariates. These findings have implications for public health measures to improve prenatal and NICU care for patients from disadvantaged areas.

**Key Points:** *Question:* Is socioeconomic deprivation at the neighborhood level, measured by an Area Deprivation Index (ADI), an independent risk factor for NICU mortality and morbidity among extremely premature infants?

*Findings:* In a cohort of 2,765 extremely premature infants (gestational age <29 weeks) admitted to four Level IV NICUs in different US regions, national ADI percentile correlated with risk of NICU mortality and morbidities after adjusting for multiple covariates.

*Meaning:* These findings have implications for public health measures to improve prenatal and NICU care for patients from disadvantaged areas.

## Introduction

Socioeconomic disadvantage impacts maternal health and access to care, affecting the preterm delivery rate ^1–3^ and infant mortality ^4^. Once premature infants leave the Neonatal Intensive Care Unit (NICU), socioeconomic deprivation in the home environment adversely impacts neurodevelopmental outcomes ^5,6^. Evidence suggests that socioeconomic disparity increases adverse outcomes of extremely preterm infants during the NICU course^7^, but the interaction between deprivation, race, and other risk factors is not well established. Racial disparities confound the effects of socioeconomic status, with non-white families bearing a higher burden of adverse health outcomes and lower quality of health care ^8–11^. In addition, due in part to a history of housing discrimination and structural racism in the US, racial minorities more often live in socio-economically disadvantaged communities^12^.

The Area Deprivation Index (ADI) is a metric that measures area disadvantage, determined by 17 Census variables, including measures of poverty, education, housing, and employment status. Singh et al. ^13^ first described this metric in relation to disparities in life expectancy that correlated with area deprivation. Kind and colleagues adapted, updated, and validated the ADI at the neighborhood level ^14^ using Block Groups, the smallest geographical unit used by the US Census Bureau. In the most recent iteration, ADI has been normalized at the national level to generate percentiles that can be easily compared, with higher values indicating greater disadvantage. National ADI percentiles and state ADI deciles using 2018 Census data are published online in an interactive map at https://www.neighborhoodatlas.medicine.wisc.edu/ ^15^.

Associations between ADI and health outcomes have been investigated in many populations, including adult patients with Alzheimer’s disease, ^16^ cardiovascular disease, ^17^ and in-hospital COVID-19 mortality ^18^. ADI has also been used to investigate disparity in neonatal health outcomes, such as the rates of exclusive breastfeeding ^19^, neurobehavioral differences and brain structure in term infants^20,21^, respiratory morbidity after NICU discharge in infants with bronchopulmonary dysplasia^22^, and length of stay in infants with neonatal opioid withdrawal syndrome ^23^.

In this project, we aimed to investigate the association of ADI, a granular, comprehensive, geospatially-linked measure of deprivation, and demographics, with in-hospital mortality of extremely premature infants and three key morbidities (late-onset sepsis, necrotizing enterocolitis and severe intraventricular hemorrhage). We hypothesized that maternal residence in areas with greater deprivation is associated with increased NICU mortality and morbidity risk. Our study includes eight years of data from four regional referral NICUs located in four US regions: Midwest, Northeast, Mid-Atlantic, and South.

## Methods

### Study design and patient population

We conducted a multicenter, retrospective cohort study of non-Hispanic Black and White premature infants born at <29 weeks’ gestation and admitted to one of four academic Level IV NICUs – University of Virginia in Charlottesville, VA, University of Alabama in Birmingham, AL, Washington University in St. Louis, MO, and Columbia University in New York City, NY – between 2012 and 2020. The maternal self-reported race was used as the infants’ race. Hispanic infants were excluded because their race was not specified in the earlier years of the study, and they accounted for relatively few patients (5-10%) at all but one NICU, Columbia University. We excluded infants with missing outcomes data from the analysis. We also excluded Asian infants and other race categories with very few patients. Each institution’s IRB reviewed and approved the study under a waiver of informed consent.

### Address geocoding and ADI data process

All geographic locations in the United States are defined using a Federal Information Processing System (FIPS) code. The degree of FIPS code granularity is determined by the number of digits, with a maximum of 15 digits to define an individual census block. Neighborhood Atlas ADI data were calculated at the US Census Block Group (BG) level, a statistical division of geographic areas containing between 600 and 3000 people. A BG is a 12-digit number: 2 digits represent the state, three digits represent the county, six digits represent the census tract, and one digit represents the block group ^24^. ADI lookup for the patient address listed in the birth encounter was performed in a semi-automated fashion using a four-step processing pipeline written in Python (See <u>Appendix</u>). Once converted, centers discarded address data, and only ADI values were shared for analysis. This way, PHI remained local and protected, and shared data were de-identified.

### Clinical and outcome data

Clinical data were extracted from unit databases or electronic health records, including demographics, perinatal variables, and outcomes. The primary outcome was mortality before NICU discharge, and secondary outcomes included late-onset sepsis or necrotizing enterocolitis and severe intraventricular hemorrhage. These secondary outcomes were defined as follows:

1. Late-onset sepsis (LOS): a positive blood culture after three days of age that was treated with antibiotics for at least five days (or less if the infant died during treatment).
2. Necrotizing enterocolitis (NEC): radiographic evidence of pneumatosis, pneumoperitoneum, or portal venous gas plus clinical signs and symptoms of sepsis according to the modified Bell’s staging criteria for stage two or higher ^25^.
3. Severe intraventricular hemorrhage (IVH): grade III-IV intraventricular hemorrhage identified on cranial ultrasound according to the Papile classification system ^26^.

### Statistical analysis

We performed univariate comparisons between ADI, demographics, and primary and secondary outcomes using Wilcoxon rank-sum or Chi-squared tests. We analyzed the multivariable relationship between ADI and NICU outcomes using Bayesian logistic regression adjusting for race, BW, outborn status, and sex. For computational efficiency, quantitative variables were centered and scaled before analysis.

In contrast with frequentist models, Bayesian models provide more information about parameters through posterior distributions ^27^, more correctly quantify the uncertainty in model parameters ^28^, and are often able to estimate models that may otherwise fail ^29^. We used weakly informative priors for all parameters. We considered a predictor significant if the posterior 95% Credible Intervals (CrI) did not include zero and took the posterior median as the summary of the posterior in all regression analyses. We compared models with interaction terms to the baseline model and considered an interaction significant if the information criterion value was lower than that of the baseline model.

## Results

### Patient characteristics

We analyzed ADI and clinical data from 2,765 extremely premature infants admitted during the study period. Of these, 13% were excluded due to missing data for multivariable modeling. Missingness occurred evenly across the four NICUs, and only 1% of infants were excluded due to missing address or ADI data. The mean gestational age was 25.6 ± 1.73 weeks, the mean BW was 805 ± 241g, 50% were male, 48% reported Black maternal race, and 18% died before NICU discharge. Table 1 summarizes cohort characteristics by center.

**Table 1.** Patient characteristics by NICU. Variables are presented as median [1st quartile, 3rd quartile] or number (%). ADI = Area Deprivation Index, UVA = University of Virginia, WUSTL = Washington University in St. Louis, CU = Columbia University, UAB = University of Alabama, NEC = necrotizing enterocolitis, Bell’s stage 2-3, Severe IVH = grade 3-4 intraventricular hemorrhage.

| Variable | UVA | WUSTL | CU | UAB |
| --- | --- | --- | --- | --- |
| N | 508 | 405 | 504 | 1348 |
| ADI national percentile | 50 [36, 66] | 75 [53 ,91] | 16 [7, 27] | 81 [64, 92] |
| Gestational Age (weeks) | 26 [24, 27] | 26 [24, 27] | 26 [24, 27] | 26 [24, 27] |
| Birth Weight (grams) | 815 [650,991] | 840 [650,1040] | 760 [624, 965] | 740 [587,950] |
| Male sex | 273 (54%) | 213 (53%) | 253 (50%) | 652 (48%) |
| Race |  |  |  |  |
| White | 371 (73%) | 230 (57%) | 252 (50%) | 587 (44%) |
| Black | 137 (27%) | 175 (43%) | 252 (50%) | 761 (57%) |
| Mortality | 68 (13%) | 72 (18%) | 81 (16%) | 277 (21%) |
| Inborn | 361 (71%) | 261 (64%) | 396 (79%) | 963 (71%) |
| Sepsis or NEC | 123 (24%) | 90 (22%) | 91 (18%) | 388 (29%) |
| Severe IVH | 71 (14%) | 94 (23%) | 26 (5%) | 162 (12%) |

### Univariate analysis

In pooled data across the four centers, the distribution of ADI was skewed, with a greater proportion of infants born to families residing in areas of higher deprivation. Mortality was similarly skewed, with more deaths in the infants with higher ADI (Figure 1) and higher ADI among non-survivors (non-survivors median [IQR] ADI 75 [45-89], survivors 64 [36-84]). Among the 1,777 (64%) infants with ADI above the national median, mortality was 20% vs. 14% in those with ADI at or below the national median (p<0.001). Higher ADI was also associated with self-reported Black maternal race, inborn status, diagnosis with sepsis or NEC, and severe IVH (Table 2). Families of Black infants lived in block groups with higher ADI than White infants (Median [IQR] ADI: Black 77 [45-93], White 57 [32, 77], Figure 3). Black infants had higher mortality (20% vs. 16%) and lower BW (785 ± 239g vs. 824 ± 241g) than White infants. We found no correlation between BW and ADI (r = −0.05). Significant inter-center variation in ADI existed, as shown in Supplemental Figure 1.

**Table 2.** Covariates and ADI distributions. Comparing ADI median [1st quartile, 3rd quartile] among infants grouped by multiple binary variables. *p<0.01 by Wilcoxon rank sum tests.

|  | Median [IQR] National ADI Percentile |  |
| --- | --- | --- |
| Died vs. Survived* | 71 [45, 89] | 64 [36, 86] |
| Black vs. White* | 77 [45, 93] | 57 [32, 77] |
| Female vs. Male | 68 [38, 87] | 64 [37, 86] |
| Inborn vs. Outborn* | 63 [35, 86] | 53 [29, 76] |
| Sepsis or NEC vs. neither* | 68 [41, 88] | 64 [35, 86] |
| Severe IVH vs. mild/no IVH* | 69 [44, 90] | 64 [36, 86] |

**Figure 1.**
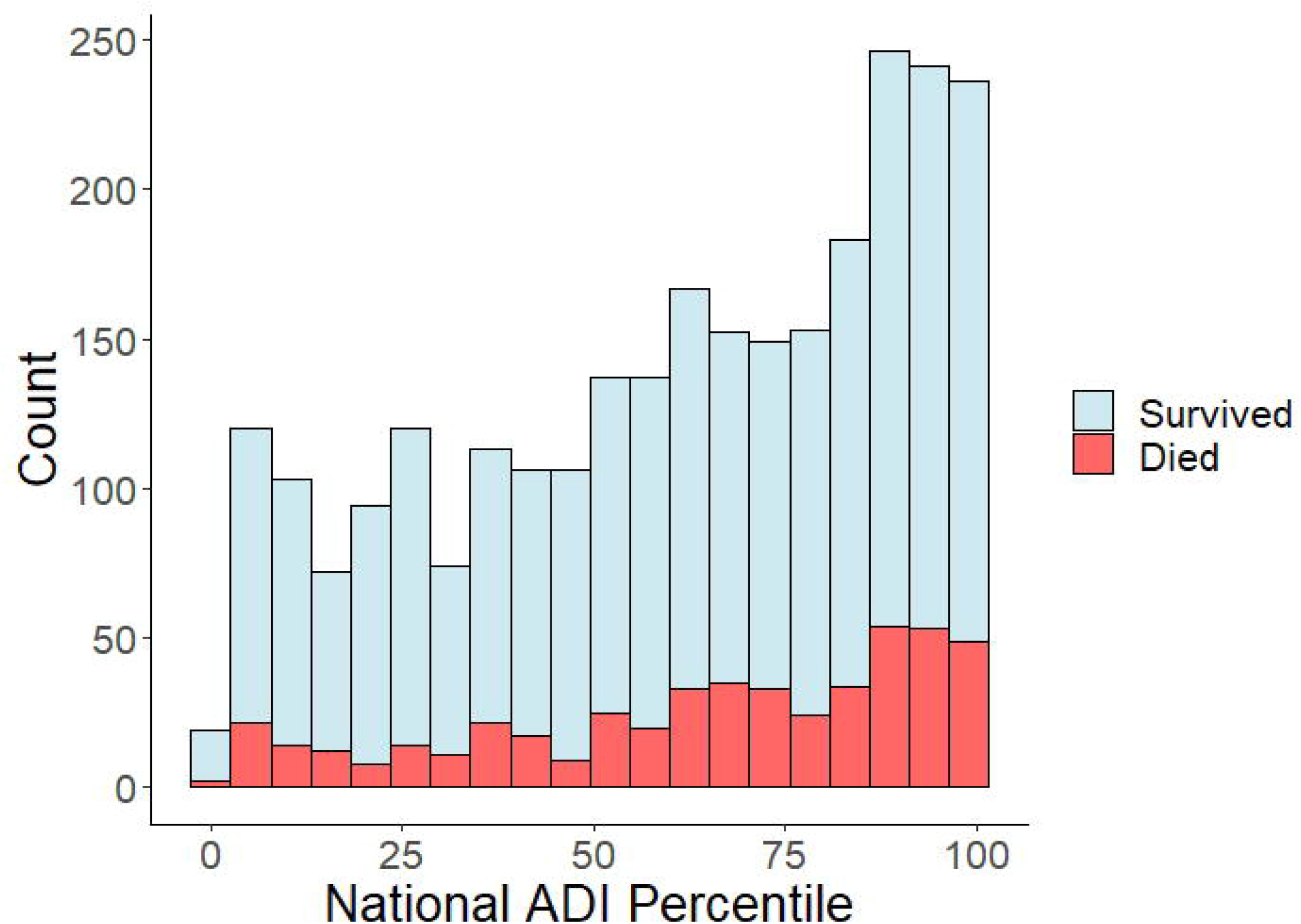
National Area Deprivation Index (ADI) and NICU mortality. The bar graph shows the number of extremely premature infants in each ADI percentile who died before NICU discharge (red) or survived (blue) on the y-axis and the ADI on the x-axis. Higher ADI indicates greater disadvantage.

**Figure 3.**
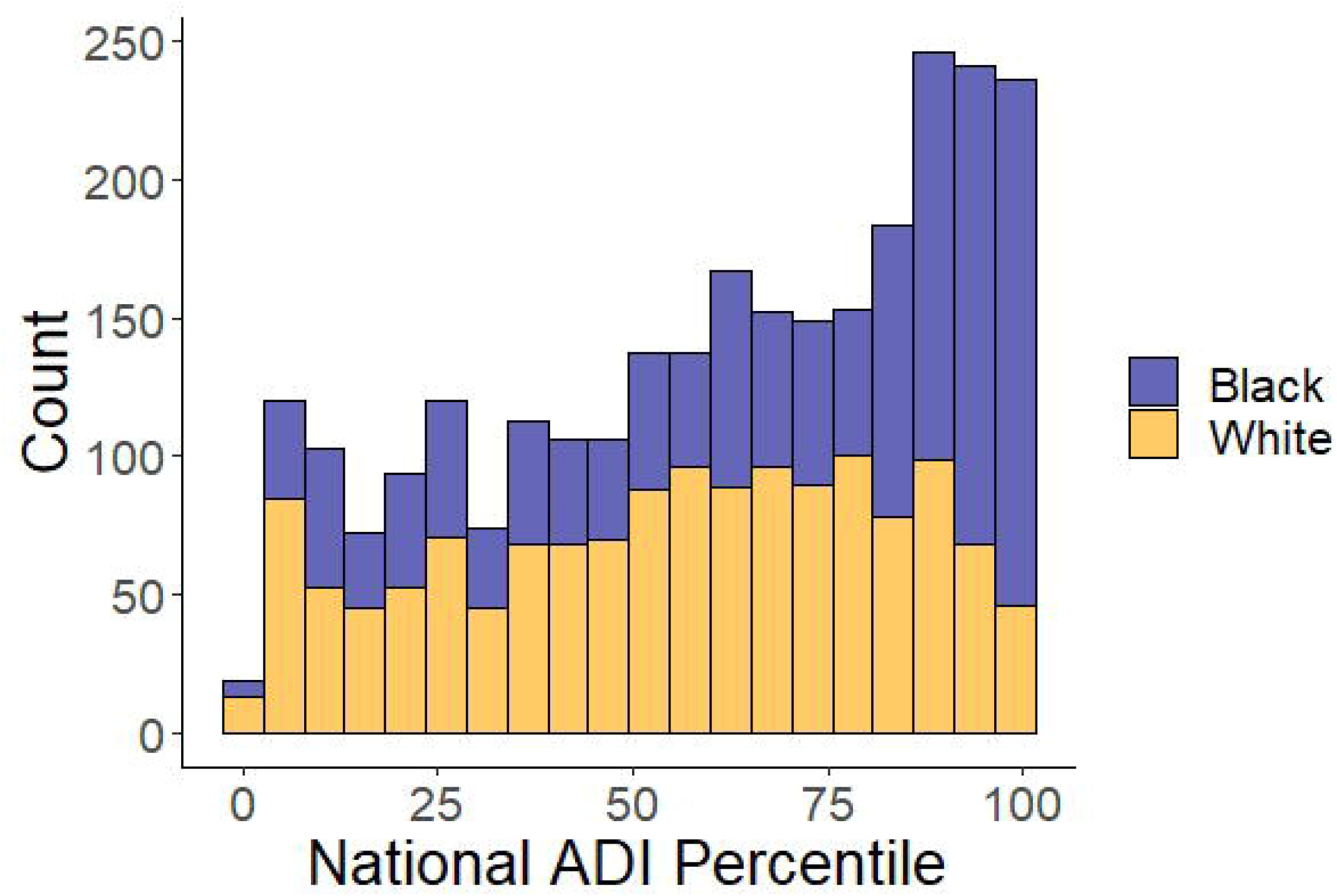
National ranked Area Deprivation Index (ADI) percentiles grouped by race. The bar graph shows the number of infants in each ADI percentile with Non-Hispanic Black or White maternal race.

Comparing male and female infants, we found no difference in ADI, the proportion of outborn infants, or mortality. As expected, the average BW was slightly higher for males than females (836 ± 243g vs. 774 ± 235g). While mortality was not significantly higher for males (18.6%) than females (17.4%) overall, White males had higher mortality than White females (17.3% vs. 15.0%), while Black infant mortality did not differ by sex (20.2% vs. 19.9%).

While most patients (72%) were admitted following delivery at the same hospital, outborn infants had lower ADI (table 2), lower mortality (15% vs. 19%), and a higher proportion of White race (64% vs. 51%). Infants transferred to the study NICU after delivery at an outside hospital also had a higher rate of severe IVH (outborn 22%, inborn 11%).

Infants with sepsis or NEC, severe IVH, or both had higher ADI than those without (Table 2, Figure 2). These morbidities were associated with higher mortality and lower BW but not with sex or race.

**Figure 2A & B.**
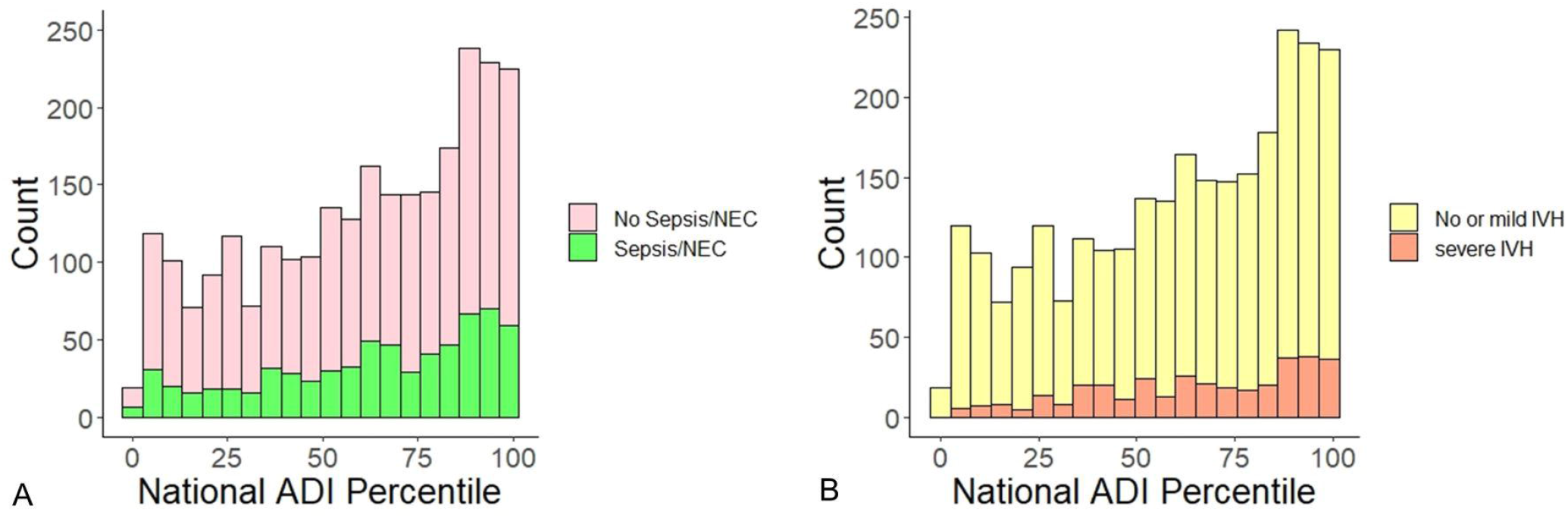
National Area Deprivation Index (ADI) percentiles and NICU morbidity. The bar graph shows the number of infants in each ADI percentile, grouped by whether or not infants had the following morbidities: Panel A) one or more episodes of confirmed sepsis or NEC; Panel B) no or mild IVH (grade 1-2) versus severe IVH (grade 3-4).

### Results of multivariable analyses

We examined the association between ADI and NICU mortality while adjusting for the covariates analyzed in univariate analyses: BW, sex, race, and outborn status. Based on posterior median estimates, BW was the strongest predictor of mortality, but national ADI percentile and male sex were also statistically significant (Table 3). Neither race nor the interaction between race and ADI was significant, and the interaction term did not improve the Watanabe-Akaike Information Criteria (WAIC) ^30^ of the model.

**Table 3.** Multivariable model results. Posterior estimates and 95% Credible Intervals (CrI) of multivariable Bayesian regression models predicting 1) NICU mortality, 2) Diagnosis of sepsis or necrotizing enterocolitis, and 3) severe intraventricular hemorrhage. Birth weight and ADI percentile were centered and scaled. Variables were considered statistically significant if the 95% CrI did not include zero (indicated in bold).

| 1. NICU Mortality |  |  |  |
| --- | --- | --- | --- |
| Predictor | Estimate | Lower 95% CrI | Upper 95% CrI |
| **Birth weight** | **-1.20** | **-1.35** | **-1.06** |
| **Male sex** | **0.35** | **0.13** | **0.57** |
| **ADI percentile** | **0.14** | **0.02** | **0.26** |
| Outborn | -0.27 | -0.58 | 0.05 |
| White race | -0.03 | -0.26 | 0.20 |
| 2. Sepsis or Necrotizing enterocolitis |  |  |  |
| Predictor | Estimate | Lower 95% CrI | Upper 95% CrI |
| **Birth weight** | **-0.44** | **-0.55** | **-0.33** |
| **Male sex** | **0.22** | **0.02** | **0.41** |
| **ADI percentile** | **0.13** | **0.04** | **0.23** |
| **Outborn** | **-0.39** | **-0.68** | **-0.12** |
| White race | -0.06 | -0.25 | 0.13 |
| 3. Severe Intraventricular Hemorrhage |  |  |  |
| Predictor | Estimate | Lower 95% CrI | Upper 95% CrI |
| **Birth weight** | **-0.33** | **-0.46** | **-0.19** |
| Male sex | 0.12 | -0.11 | 0.36 |
| **ADI percentile** | **0.27** | **0.14** | **0.40** |
| **Outborn** | **0.93** | **0.65** | **1.21** |
| White race | 0.11 | -0.14 | 0.37 |

BW and ADI were also significant predictors in multivariable models of NICU morbidities (Table 3). Because we scaled and centered continuous variables, we were able to compare the size of the estimates among variables included in each model. By treating ADI and BW values as Z scores, we report the effect of changing either variable by one standard deviation (SD), rather than by one percentile (ADI) or one gram (BW). Therefore, in the model for mortality, we interpreted the effect of decreasing BW by one SD to be nine times that of increasing ADI by one SD. In contrast, in the model for severe IVH, the effects of BW and ADI were similar and still less than the effect of outborn status. When modeling sepsis and NEC, the effect of decreasing BW was about three times that of increasing ADI, and male sex and outborn status had intermediate effects. Race did not significantly affect any of the three outcomes modeled.

## Discussion

Socioeconomic factors play an important role in maternal health, access to prenatal care, rates of preterm birth, and infant morbidity and mortality outside of the NICU ^4,5,31^. While in the NICU, preterm infants have standardized care guidelines that might be expected to limit the impact of socioeconomic deprivation on adverse outcomes. Few studies have evaluated the effect of area deprivation on in-hospital outcomes^32^. In this study, we found that residence in a disadvantaged neighborhood- as measured by the ADI - increases the risk of an extremely preterm infant’s dying or developing morbidity, even after accounting for risk factors. This indicates a pervasive and detrimental effect of area deprivation that may not be entirely eliminated by provision of standardized, high-quality NICU care.

The mechanism by which area deprivation affects neonatal outcomes may include the effect of maternal adversity on the developing fetus ^33,34^. Maternal stress and poor access to prenatal care have been linked with adverse birth outcomes ^35–37^. Lower maternal socioeconomic status increases the risk of preterm birth ^11,38,39^ and among those born prematurely, it increases the risk of adverse neurodevelopmental outcomes^40,41^. In contrast with other studies, we did not find an association between ADI and BW, likely due to the fact that we limited the analysis to extremely premature infants with a narrow range of BW.

We found that the association between higher deprivation and mortality remained significant in multivariable analysis while the association between Black race and mortality did not. These highly collinear variables make it difficult to estimate the effect of race or deprivation alone. Studies indicate that despite advances over time, racial disparities persist in neonatal outcomes and care practices, ^10,42,43^ but these analyses did not account for socioeconomic factors. As non-White race groups endure more socioeconomic deprivation, it becomes essential to consider social disparities as a significant mediator of racial disparities. Public health efforts have been made to narrow disparities in antenatal steroid usage and cesarean deliveries, but gaps in these practices persist ^43^. Disparities in breastmilk utilization by race persist ^10^ despite overall improvements in breastfeeding initiation over time, with the lowest rate of breastmilk feeding at NICU discharge among infants born to non-Hispanic Black mothers ^44^. In our cohort, infants born to Black mothers were exposed to higher levels of deprivation in-utero and had lower BW. Still, they did not have higher associated morbidity or mortality when accounting for these and other confounding variables. Our results suggest that the higher mortality observed among Black preterm infants compared with White may be partly confounded by factors captured in the ADI, including exposure to adverse social and economic conditions.

A strength of this study was the inclusion of four centers in geographically distinct areas (Northeast, Mid-Atlantic, Midwest, South) with different distributions of ADI, patient racial compositions, and population densities. Although all centers are level IV referral NICUs, there were differences in the percentage of inborn infants, ranging from 64 to 79%. Interestingly, the distribution of ADI was skewed in opposite directions for infants admitted to the Midwestern and Southern centers compared with the Northeastern center, while the Mid-Atlantic center had a nearly symmetric ADI distribution (Supplemental Figure 1). The distributions within each center cohort appear to be representative of the regional distributions displayed on the Neighborhood Atlas ADI map. We did not adjust for center in multivariable modeling as national percentile ADI data has already been normalized across all geographic regions. Additionally, we did not use state ADI deciles in our analysis since these values do not translate across state lines and would not be appropriate for a pooled sample including data from multiple states.

Limitations of the study include the lack of granularity in the ADI and race data. Although block groups are the most geospatially compact census unit and the ADI can provide a localized overview of the degree of socioeconomic deprivation, its value is opaque. The relative contributions of each ADI component are not available for the most current values, making it infeasible to evaluate driving factors of the association between ADI and NICU morbidity and mortality. In addition, the exposures captured in the ADI may have differential impacts across the heterogeneous locations that we analyzed. For example, in the US 2020 Census data for just the four cities in which the study NICUs were located, the land area ranged between 10.24 and 302.64 mi^2^; population density ranged between 1,365 and 29,302 per mi^2^; median household incomes ranged between $38,832 and $67,046, and the percentage of adults without health insurance ranged between 7.9 and 14.7%. Each of these differences likely plays a significant role in the factors which underlie the ADI calculation.

We classified race into broad groups of Black or White based on the self-reported maternal race and excluded Hispanic, Asian, and American Indian and Alaska Native patients, which may have left important interactions or effects undiscovered. Analyzing more refined racial and ethnic subgroups, including mixed race when the father’s race differs from the mother’s race, may have found differential associations between ADI and outcomes among more finely delineated racial and ethnic groups. For example, the “Hispanic Paradox” refers to the paradoxical finding that Hispanic infant mortality rates lie at or below the rate of Non-Hispanic White or Caucasian infant mortality despite higher socioeconomic deprivation ^45,46^.

Our results generate questions for future studies to evaluate how interventions to reduce area deprivation or improve equity might reduce disparities in NICU outcomes for premature infants. The study’s findings also leave us wanting to know more about the mechanism of the association between ADI and NICU outcomes, but we could not answer such questions with the data available. Future analyses should investigate the ADI and its individual components as mediators or moderators of adverse outcomes in premature infants.

## Conclusions

Among preterm infants <29 weeks’ gestation admitted to four NICUs, ADI was a risk factor of NICU mortality and morbidity after adjusting for multiple covariates. These findings highlight the need for further research on the components of the ADI that drive this association. Specific risk factors may be targets of public health measures to improve prenatal and NICU care for patients from disadvantaged areas.

## Data Availability

All data produced in the present study are available upon reasonable request to the authors

https://github.com/AyushDoshi/geocode-adi

