## Appendix for "Neighborhood deprivation is associated with NICU mortality for extremely premature infants: A 4-NICU study"

Supplemental Material

ADI data processing pipeline

1. The address list was first converted to a Pandas DataFrame.
2. The corresponding block group (BG) was then identified for each patient. Given the complexities of postal addresses, four sub-steps were needed to account for the range of nuances.
   i. P.O. and rural routes were removed from the address list and set aside
   ii. First-pass conversion to BG was attempted by entering addresses directly into the US Census Geocoder (https://geocoding.geo.census.gov/geocoder/)
   iii. For addresses that failed geocoding, a second-pass conversion was attempted using the latitude and longitude of the address in decimal degree format. Coordinates were obtained using the Google Maps API or the Nominatim API (https://nominatim.org/), which utilizes the OpenStreetMap database.
   iv. Addresses successfully converted to latitude/longitude coordinates were re-entered into the US Census Block Group Geocoder to obtain the BG.
3. For all addresses where a BG could be identified, the corresponding national percentile and state decile ADI were mapped by BG and exported to a results file. The ADI data were developed by researchers at the University of Wisconsin from 2018 Census data (Kind and Buckingham 2018).
4. Addresses that could not be converted to a BG were then manually reviewed. In many cases, the address contained a typographical error or was partially incomplete and could be re-run through the script. For PO boxes and rural routes, the BG of the corresponding Post Office was used. Patients with significant address errors, missing data, or false addresses were excluded.

Code is available in a GitHub repository (https://github.com/AyushDoshi/geocode-adi).

Supplemental Figures

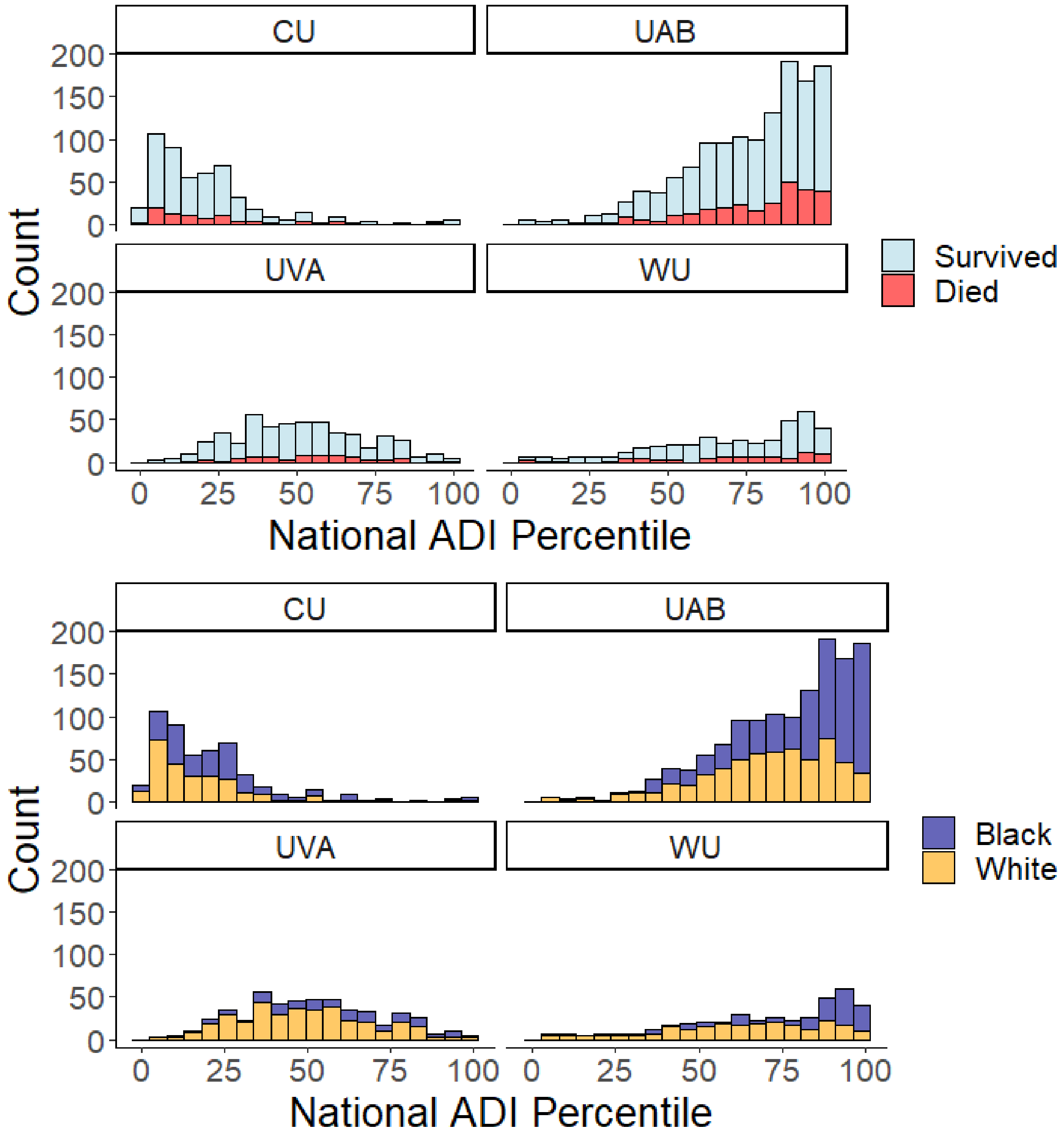


Supplemental Figure 1. ADI histograms grouped by mortality (Top Panel) and race (Bottom Panel) for infants at each of the four NICUs. UVA = University of Virginia, WU = Washington University in St. Louis, CU = Columbia University, UAB = University of Alabama. Higher ADI indicates greater deprivation.
